# Field evaluation of Standard Q Filariasis Antigen Test for Lymphatic Filariasis (LF) during a pre-transmission assessment survey in Sierra Leone, 2022

**DOI:** 10.1101/2024.12.12.24318905

**Authors:** Benoit Dembele, Mohamed Salieu Bah, Abdulai Conteh, Habib Kamara, Ibrahim Kargbo-Labour, Ashley Souza, Patricia Houck, Ernest O. Mensah, Victoria Turay, E. Scott Elder, Katherine Gass, Steven D Reid, Joseph P. Shott, Yaobi Zhang, Kimberly Y. Won, Angela Weaver

## Abstract

**Background:** As part of a multi-country evaluation, the SD Biosensor STANDARD™ Q Filariasis Ag Test (QFAT) was compared with the Abbott Bioline TM Filariasis Test Strip (FTS) for classifying lymphatic filariasis (LF) prevalence at a population level and for ease of use in field conditions in Sierra Leone.

**Methods and principal findings:** The evaluation was done in two districts, Bombali and Karene, where repeat pre-transmission assessment surveys (pre-TAS) were planned. Two sites with high LF antigen prevalence in 2020 (4.1% in the village of Kagbo and 7.7% in the village of Makorba Yelimi) were chosen. Convenience sampling was used to recruit 350 community members ≥5 years in each site. Blood was collected by fingerstick (20μl for QFAT and 75 μl for FTS). The reading time for both tests was 10 minutes. For all positive or invalid results, a repeat test was performed for both tests. In total, 728 participants (5 - 91 years) were tested by QFAT and FTS. The positive rate was 4.8% (17/357) and 3.5% (13/367) for FTS and 3.4% (12/357) and 4.1% (15/367) for QFAT in Kagbo and Makorba Yelimi, respectively. All participants testing positive for FTS or QFAT underwent further testing by night blood smear to detect microfilariae using microscopy. None of the positive participants had circulating microfilariae. Nearly half (14/30) of those who tested positive with FTS during this survey also tested positive with FTS in re-pre-TAS in 2020. Four FTS and three QFAT samples were indeterminate (meaning a positive result followed by a negative result). In field conditions, QFAT was easy to handle and recorded zero invalid tests compared to FTS (six invalids). Using the FTS results as a reference standard, the sensitivity and specificity of the QFAT was 78.6% and 99.4% respectively. The concordance between FTS and QFAT was 0.81 (Cohen’s Kappa). The discrepancy found between the two tests in terms of positive tests was not statistically significant (p=0.78).

**Conclusions / significance:** The results suggest that the QFAT is a credible LF diagnostic test when compared to the routinely used FTS; use of either test would result in the same program decision.

**Authors summary:** Lymphatic filariasis (LF) is a vector-borne disease targeted for elimination as a public health problem by 2030. The Global Program to Eliminate LF recommends tools to measure the impact of interventions and to achieve elimination. A reliable and easy to use diagnostic tool is key for the success of the global program. Currently only one rapid antigen test is used for programs in *Wuchereria bancrofti* endemic counties. This study was part of a multi-country field evaluation of the SD Biosensor STANDARD™ Q lateral flow assay rapid diagnostic test. The primary objective was to determine comparability of the SD Biosensor STANDARD™ Q Filariasis Ag Test (QFAT) to the Abbott Bioline™ Filariasis Test Strip (FTS) in its ability to classify LF prevalence at a population level. In addition, information was collected on the utility and ease of use of the QFAT in field settings. The evaluation was done in two districts (Bombali and Karene) in Serra Leone, which were undergoing repeat pre-transmission assessment surveys (pre-TAS). The results of this study confirm the performance of QFAT as a suitable alternative to the currently recommended FTS. In field conditions, using QFAT seems effective given that it records zero invalid tests compared to the FTS (six invalid tests).

## Introduction

Lymphatic filariasis (LF) is a preventable mosquito-borne infectious disease, caused by infection with one of the filarial parasites, *Wuchereria bancrofti, Brugia malayi* and *Brugia timori*. The infection impairs the lymphatic system, where the worms nest, later manifesting in some infected individuals as hydrocoele, lymphoedema and elephantiasis, resulting in unnecessary physical and mental suffering [1]. *W. bancrofti* is found in nearly all LF endemic countries and *Brugia* spp. are found only in limited areas of a few countries across Southeast Asia [2]. *W. bancrofti* is responsible for 90% of the cases of LF [3]. The World Health Organization (WHO) established the Global Program to Eliminate Lymphatic Filariasis (GPELF) to eliminate LF by mass drug administration (MDA) of anthelminthics and to alleviate the suffering of people affected by the disease through morbidity management and disability prevention (MMDP). According to the WHO 2021–2030 road map for neglected tropical diseases (NTDs), the global goal for LF is elimination as a public health problem by 2030 through repeated annual rounds of MDA [4]. By 2022, 19 of the 72 endemic countries had successfully eliminated LF as a public health problem. However, there are still an estimated 794 million people requiring MDA worldwide [1]. To achieve LF elimination, a country follows four recommended sequential programmatic steps: baseline mapping, MDA implementation, post-MDA surveillance, and validation of LF elimination. All four stages of the program are dependent on the availability of user-friendly and highly sensitive and specific rapid diagnostic tools [5].

Where *W. bancrofti* is the causative agent of LF, the Alere Filariasis Test Strip (FTS, Abbott) is currently the main diagnostic tool recommended for program use [2]. The FTS detects circulating filarial antigen (CFA), indicating the presence of an adult worm. However, FTS has limitations. First, it requires sufficient time to collect 75 microliters (μL) of blood using micropipettes, which increases the risk of clotting, preventing proper flow of the blood sample on the test strip. Second, FTS consists of a single lightweight nitrocellulose strip devoid of any protective housing (e.g., a plastic cassette) and a lightweight plastic container, which has created logistical challenges, requiring survey teams to secure the test strips with tape to minimize movement during testing.

A new rapid diagnostic test, the STANDARD™ Q Filariasis Antigenemia Test (QFAT) (SD Biosensor, Suwon, South Korea) also detects CFA and may be an alternative tool to the FTS. The test cassette form factor required small sample volume (20 μL) and inclusion of a buffer to aid in sample flow represent potential improvements compared to the FTS. We previously conducted a multi-country field evaluation of the QFAT in four countries in the Pacific [6], Asia and Africa regions. Here, we report comparison of the QFAT to FTS in Sierra Leone.

Previously, Sierra Leone was endemic for LF throughout the country [7]. LF MDA started in Sierra Leone in 2007, achieving 100% geographical coverage in 2009 [8, 9]. Despite various challenges, the country has achieved great progress toward LF elimination [8, 10, 11]. To date, 12 of 16 districts have stopped LF MDA and transitioned to post-MDA surveillance [12]. Four districts (Bombali, Karene, Koinadugu, and Falaba) remained under MDA and were scheduled to conduct a fourth repeated-pre-transmission assessment survey (pre-TAS) using FTS in 2022, in line with the recommendations in the WHO guideline [12] [13]. Due to the inclusion of CFA detection in the survey, this provided a good opportunity for a comparative evaluation of QFAT and FTS.

## Methods

### Ethical Considerations

The survey protocol was approved by the Ethics and Scientific Committee, Ministry of Health and Sanitation, Sierra Leone. Consent documents and participation information were provided and explained by team members who were fluent in the appropriate local language to participants. The community leaders were informed about the survey and their permission was sought and granted to conduct the survey in their respective communities. Before sample collection, participants were sensitized to the nature of the survey, and individual consent forms for adults and assent for minors (5 - 17 years) were obtained. Participants were free to leave at any time during the survey. To ensure privacy and confidentiality, an identification barcode was assigned to each participant and used only for sample tracking. Personal identifying information was kept confidential and is not included in this report. All confirmed positive and indeterminate cases were treated with ivermectin and albendazole based on the survey protocol by the survey team.

### Study sites

The study was conducted in two villages in Sierra Leone: Kagbo village in the Bombali district and Makorba Yelimi village in the Karene district from July 18– 22, 2022. These two villages were purposively selected for this study. They were included and surveyed as spot check sites in the last failed pre-TAS in 2020. The Kagbo village is located in the Safroko Limba chiefdom in Bombali and had an antigenemia prevalence by FTS of 4.1% in 2020. The Makorba Yelimi village is located in the Sanda Loko chiefdom and had an antigenemia prevalence of FTS of 7.7% in 2020.

### Study population, sampling and data collection

During the survey, the team arrived at each community early to brief the stakeholders and set-up enrollment and data collection points. Community members ≥18 years provided written or fingerprint informed consent; parental permission was received for participants 5-17 years. Participants were enrolled by convenience sampling, the standard sampling method for pre-TAS. Demographic data (name, age, sex) were collected, and information on the length of stay in the district and participation in previous MDAs was documented. Each participant was assigned a unique identification number using a QR code, which was used to label the consent form, all the samples collected, and the tests for the same person. Global positioning system (GPS) coordinates were recorded for each survey site. Standardized questionnaires were used to collect the team technicians’ opinions on the ease-of-use of QFAT. All data were entered into an electronic form using Open Data Kit (ODK) and ONA platform installed on tablets.

### Diagnostic procedures

Prior to the survey, two tests from each batch of FTS were tested with a positive control obtained from WHO at the Central Public Health Laboratory in Freetown. For the QFAT, quality control testing using the same positive control test was carried out during the training of the survey team. All test kits were stored at ambient temperature (16°-24°C) in the laboratory prior to the survey. Sample collection protocols differed between the two study sites. In Kagbo village, two technicians were involved in blood collection and conducting the tests. First, the technicians pricked the finger of a participant using a sterile disposable lancet. A 20µl blood sample was collected directly from the finger prick using a sample collection device included with the QFAT test. The blood sample was placed on the cassette sample port, and two drops of buffer solution were added. Then, approximately 190µl of finger prick blood was collected into a heparinized microtainer tube (Ram Scientific) according to the standard operating procedures established for the project. Subsequently, 75µl of blood was taken from the heparin tube using a pipette and added slowly to the lower half of the sample pad of the FTS. QFAT and FTS results were read at exactly 10 minutes (recorded by digital timers) according to manufacturer instructions. The remaining blood sample in the heparinized microtainer tube was placed on the six extensions (10µl each) of a filter paper disc (Trop Bio) to prepare dried blood spot (DBS) samples. In Makoba Yelimi village, blood samples were directly collected from the finger prick using the manufacturer provided sample collection devices for the FTS (75µl) and QFAT (20µl). Tests were performed according to manufacturer instructions. DBS samples were then collected directly from the finger.

All positive results for FTS or QFAT were repeated with a second FTS or QFAT test using the blood sample collected and stored in the heparin tube in Kagbo or a fresh blood sample collected from a finger prick in Makoba Yelimi. Only invalid (i.e. absence of control line or failure of complete sample migration on test strip) FTS, not QFAT, tests were repeated with a second test. A test result for FTS or QFAT was considered positive when the first and the second tests were positive. The test result was considered indeterminate when the first test was positive and the second test was negative, or both tests were invalid.

Individuals who were confirmed positive by FTS or QFAT were followed up for a night blood specimen collection during the hours of peak microfilariae (Mf) circulation (10 pm to 2 am). Slides were labeled with the corresponding QR codes of the individuals. A blood sample was collected directly from the finger prick and gently placed on a slide as three lines along the length of the slide giving a total of 60μl of finger prick blood for each slide (3×20µl). Two slides were prepared per individual. The slides were transported to the Neglected Tropical Disease laboratory in Makeni where they were stained with Giemsa and examined for the presence of Mf.

### Ease of use questionnaire

Survey technicians who implemented the tests were interviewed to get their opinion on QFAT only. An electronic questionnaire was sent to the users to grade (strongly disagree, disagree, agree or strongly agree) the questions on the QFAT instructions for use, kit packaging and labeling, kit packing material, procedures, interpretation of results.

### Data Analysis

Data from the ODK website were exported into Microsoft Excel, cleaned and analyzed using Statistical Package for Social Science (SPSS version 20) and Epi info 7. Data were analyzed by gender and age group (5-9, 10-14, 15-19, ≥ 20 years). Site prevalence by FTS or QFAT was estimated as percentage of number of positive cases to the number of valid tests for each test. Chi-squared test was used to compare proportions between the two tests, and the Cohen’s Kappa coefficient, a measure of agreement between the two tests was used for concordance. The responses to the ease-of-use questionnaire were tallied and each reported response was converted to a value of 1.

## Results

### Prevalence

In total, 728 participants aged 5 - 91 years, 383 females (52.60%) and 345 males (47.39%), were tested by both QFAT and FTS. The proportion of the participants by age group is shown in Table 1. Overall, 96.84% (705/728) of participants recalled swallowing ivermectin and albendazole during the last MDA. Among the 23 participants who did not swallow the drugs, 52.17% (12/23) reported that they were not offered the MDA drugs because they were absent or unaware of the MDA. Also, 0.96% (7/728) participants had lived in the community less than one year prior to the survey whilst 85.16% had lived in the community since they were born.

**Table 1.**
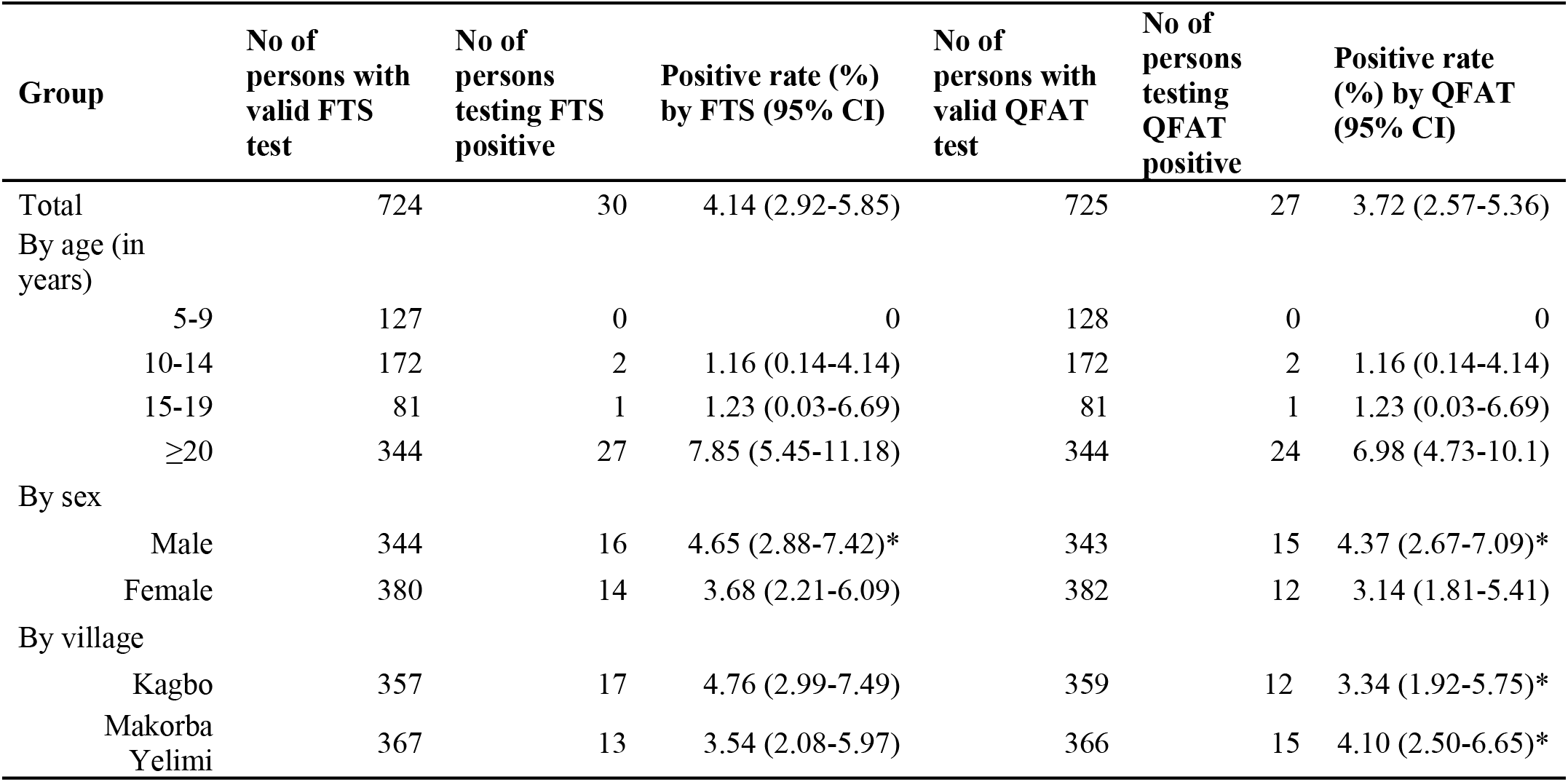
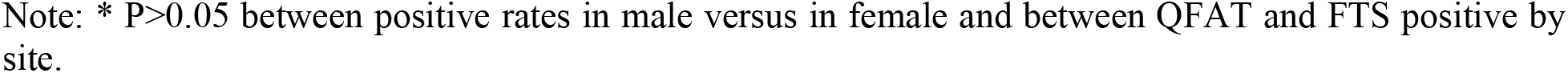
FTS and QFAT result distribution by age group.

| Group | No of persons with valid FTS test | No of persons testing FTS positive | Positive rate (%) by FTS (95% CI) | No of persons with valid QFAT test | No of persons testing QFAT positive | Positive rate (%) by QFAT (95% CI) |
| --- | --- | --- | --- | --- | --- | --- |
| Total | 724 | 30 | 4.14 (2.92-5.85) | 725 | 27 | 3.72 (2.57-5.36) |
| By age (in years) |  |  |  |  |  |  |
| 5-9 | 127 | 0 | 0 | 128 | 0 | 0 |
| 10-14 | 172 | 2 | 1.16 (0.14-4.14) | 172 | 2 | 1.16 (0.14-4.14) |
| 15-19 | 81 | 1 | 1.23 (0.03-6.69) | 81 | 1 | 1.23 (0.03-6.69) |
| $\geq 20$ | 344 | 27 | 7.85 (5.45-11.18) | 344 | 24 | 6.98 (4.73-10.1) |
| By sex |  |  |  |  |  |  |
| Male | 344 | 16 | 4.65 (2.88-7.42)* | 343 | 15 | 4.37 (2.67-7.09)* |
| Female | 380 | 14 | 3.68 (2.21-6.09) | 382 | 12 | 3.14 (1.81-5.41) |
| By village |  |  |  |  |  |  |
| Kagbo | 357 | 17 | 4.76 (2.99-7.49) | 359 | 12 | 3.34 (1.92-5.75)* |
| Makorba Yelimi | 367 | 13 | 3.54 (2.08-5.97) | 366 | 15 | 4.10 (2.50-6.65)* |

Note: \* $P > 0.05$ between positive rates in male versus in female and between QFAT and FTS positive by site.

Of 728 participants tested by both QFAT and FTS, 4.14% (30/724) were antigen positive by FTS, whereas 3.72% (27/725) were positive by QFAT. There was no statistically significant difference between the results of the two tests (χ^2^=0.08, p=0.78). The positive rate was 4.76% (17/357) and 3.54% (13/367) for FTS and 3.34% (12/359) and 4.10% (15/366) for QFAT in Kagbo and Makorba Yelimi villages, respectively, however the difference between the two tests was not statistically significant (χ^2^=0.60, p=0.43 and χ^2^=0.04, p=0.84, respectively). None of the positive participants had circulating Mf from the midnight blood tests. Nearly half (14/30 and 13/30) of those who tested positive respectively with FTS and QFAT during this survey also tested positive with FTS during the re-pre-TAS in 2020. More positive cases were recorded among those ≥20 years regardless of the type of test kit used; 7.85% (27/344) and 6.98% (24/344) for FTS and QFAT respectively. No positive case was found in the age group of 5 to 9 years old for either test. More males than females were positive by both tests, but the difference was not statistically significant (for FTS (χ^2^= 0.22 p=0.64) and QFAT (χ^2^= 0.46 p=0.50)). A total of seven tests were indeterminate: four for FTS and three for QFAT.

### Concordance

**Table 2.** Concordance of results between two tests.

|  | FTS positive | FTS negative | FTS Indeterminate | Total |
| --- | --- | --- | --- | --- |
| QFAT positive | 22 | 4 | 1 | 27 |
| QFAT negative | 6 | 689 | 3 | 698 |
| QFAT Indeterminate | 2 | 1 | 0 | 3 |
| Total | 30 | 694 | 4 | 728 |

The concordance test between FTS and QFAT, Cohen’s Kappa was 0.81 indicating strong agreement between tests. There were six invalid tests for FTS, but none for QFAT. Five FTS-invalid tests occurred in the first tests, while one occurred in the confirmatory test. Two of the six FTS-invalid tests were QFAT positive.

### User experience

The test users (5 interviews) generally agreed in their responses to the ease-of-use questionnaire. The questions were related to instruction for use, kit packaging and labelling, kit packing material, Device Assay procedure, interpretation of result (Table 3).

**Table 3.**
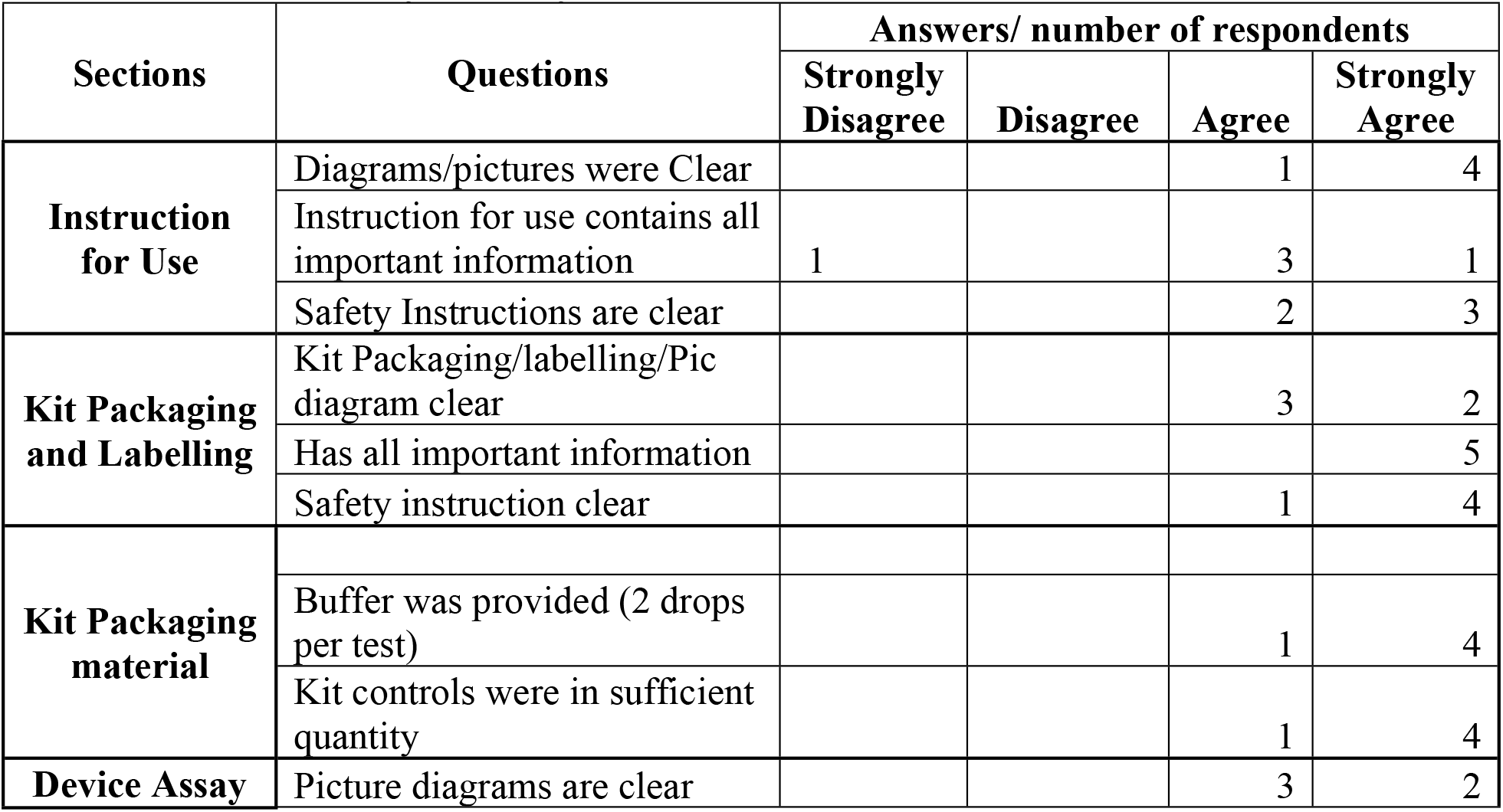
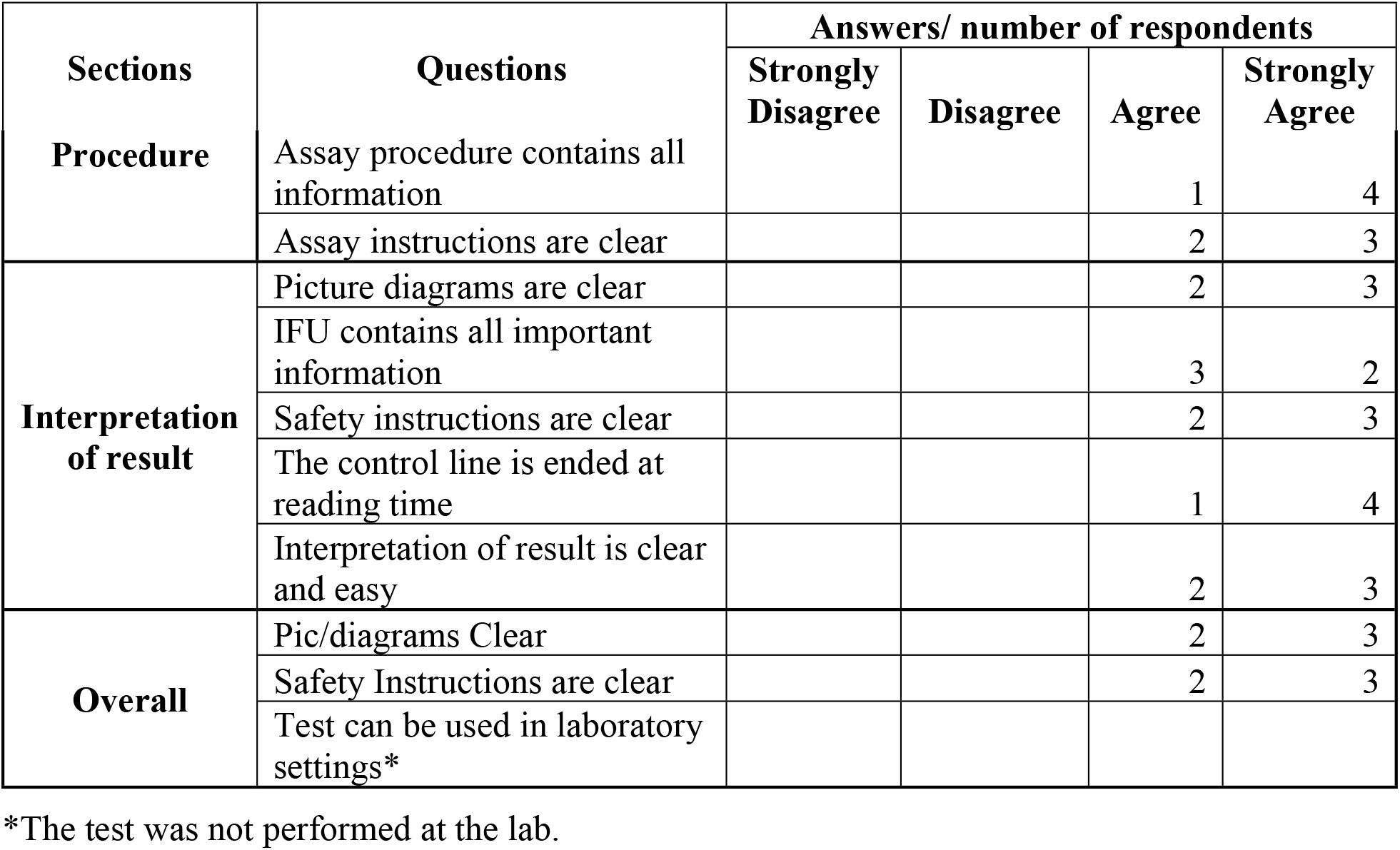
QFAT user survey summary.

| Sections | Questions | Answers/ number of respondents |  |  |  |
| --- | --- | --- | --- | --- | --- |
|  |  | Strongly Disagree | Disagree | Agree | Strongly Agree |
| **Instruction for Use** | Diagrams/pictures were Clear |  |  | 1 | 4 |
|  | Instruction for use contains all important information | 1 |  | 3 | 1 |
|  | Safety Instructions are clear |  |  | 2 | 3 |
| **Kit Packaging and Labelling** | Kit Packaging/labelling/Pic diagram clear |  |  | 3 | 2 |
|  | Has all important information |  |  |  | 5 |
|  | Safety instruction clear |  |  | 1 | 4 |
| **Kit Packaging material** |  |  |  |  |  |
|  | Buffer was provided (2 drops per test) |  |  | 1 | 4 |
|  | Kit controls were in sufficient quantity |  |  | 1 | 4 |
| **Device Assay** | Picture diagrams are clear |  |  | 3 | 2 |
|  |  | Strongly Disagree | Disagree | Agree | Strongly Agree |
| Procedure | Assay procedure contains all information |  |  | 1 | 4 |
|  | Assay instructions are clear |  |  | 2 | 3 |
| Interpretation of result | Picture diagrams are clear |  |  | 2 | 3 |
|  | IFU contains all important information |  |  | 3 | 2 |
|  | Safety instructions are clear |  |  | 2 | 3 |
|  | The control line is ended at reading time |  |  | 1 | 4 |
|  | Interpretation of result is clear and easy |  |  | 2 | 3 |
| Overall | Pic/diagrams Clear |  |  | 2 | 3 |
|  | Safety Instructions are clear |  |  | 2 | 3 |
|  | Test can be used in laboratory settings* |  |  |  |  |

\*The test was not performed at the lab.

## Discussion

The study in two villages in Sierra Leone was among the first field testing of QFAT to detect LF antigenemia, comparing its performance with FTS. The prevalence of LF by QFAT and FTS at both sites was comparable (χ^2^=0.08, p =0.78). Both villages recorded filarial antigen prevalence of >2% by either FTS or QFAT test, indicating that the same program decision (failing to pass the re-pre-TAS) was made based on the results of either test. The prevalence of LF Ag was comparable by sex 4.65% (16/344) in males versus 3.68% (14/380) in females for FTS and 4.47% (15/343) in males versus 3.14% (12/382) in females for QFAT, respectively. No statistical difference was observed between the two tests by site; the positive rate was 4.8% (17/357) and 3.5% (13/367) for FTS and 3.4% (12/359) and 4.1% (15/366) for QFAT in Kagbo and Makorba Yelimi villages, respectively. The Cohen’s Kappa coefficient, a measure of agreement between the two tests, was 0.81, suggesting a strong concordance. These metrics indicate that the QFAT performs reliably as a valid diagnostic tool for LF in field conditions, as compared to the performance of the FTS. This result corroborates that of the QFAT/FTS comparison study carried out in a field laboratory in Samoa, where the concordance rate was 0.85 [6]. Laboratory comparisons of QFAT and FTS using serum or plasma samples showed that the QFAT was a suitable rapid antigen test for use in LF elimination programs and had advantages over FTS in ease-of-use, smaller sample, clearer control line, and higher sensitivity for Mf-positive samples [14].

One of the primary goals of the study was to assess the practicality of the QFAT in field conditions. The fact that the quantity of blood required for the QFAT is smaller (20 µl) than for the FTS (75 µl) makes it less challenging for surveyors in the field to collect a sufficient volume of blood for the test. The ease-of-use questionnaire responses indicated generally strong agreement that the QFAT had good instructions and was well packaged. In addition, the QFAT test cassettes can easily be directly labelled with ID numbers, unlike FTS, which need to be fixed to the test reservoirs first before adding blood, a step not required for QFAT. While the QFAT requires an additional step of adding a buffer that is not required with the FTS, the inclusion of the buffer facilitated the migration of the blood sample and high throughput, while poor migration is often observed with the FTS. The sample collection device for QFAT was easy and more convenient for blood collection than the FTS device. More importantly, the difference in shelf life (12 months for FTS vs. 24 months for QFAT) has important implications for program use.

While we did not directly assess the test line intensity in our qualitative evaluations, the end users noted that the QFAT test line was sometimes faint and thin, possibly making it challenging to interpret positive results accurately. This was one of the noted disadvantages of the QFAT in this study. This may lead to difficulties in field conditions where clarity and ease of result interpretation are crucial for timely and accurate decision-making.

To improve the QFAT and address the issue of thin test lines, it is suggested that developers focus on several key areas. They could enhance the line visibility by increasing the contrast and thickness of the positive test line to make results more pronounced and easier to interpret. Providing comprehensive training and clear guidelines for interpreting test results is also crucial. Lastly, implementing robust quality assurance programs is essential to ensure that test kits meet high standards and perform consistently across different settings. Quality assurance programs, as highlighted by Gass et al. (2012) [15], are critical for maintaining the reliability of diagnostic tests in field conditions. These combined efforts may enhance the reliability of the QFAT, making it an effective tool for diagnosing LF. The QFAT is an opportunity for Global LF elimination program to have a second reliable antigen test to sustain LF survey implementation with less issue around the test’s availability.

## Conclusions

Overall, the QFAT’s performance in terms of concordance with the FTS suggests that it is a reliable diagnostic test for LF. Its ease of use, high throughput, and zero invalid test results in field conditions further enhance its practicality. However, more investigations are required in different prevalence settings to confirm that the QFAT is a reliable alternative for the FTS. Overall, this field evaluation of the QFAT in a program setting demonstrated comparable performance to the FTS and greater ease of use than FTS in field conditions. Our results suggest that QFAT can be used in GPELF as an alternative to the FTS.

## Data Availability

The data are available in the manuscritp in tables.

## Acknowledgements

The authors thank the WHO Diagnostic Technical Advisory Group LF subgroup and SD Biosensor for providing the tests. A special thank you to Jonathan King (WHO Geneva) for all his technical support and guidance for data analysis. A huge thank you to the survey implementing team and the communities in Sierra Leone where the survey took place.

## Funding

This work received financial support from the Coalition for Operational Research on Neglected Tropical Diseases (COR-NTD), which is funded at The Task Force for Global Health primarily by the Bill & Melinda Gates Foundation and the United States Agency for International Development (USAID) through its Neglected Tropical Diseases Program (grant OAA-13-000008). It was partially supported by USAID’s Act to End Neglected Tropical Diseases | West Program (cooperative agreement #7200AA18CA00011) managed by FHI 360 and implemented by Helen Keller Intl in Sierra Leone. JPS is funded by USAID.

## Disclaimer

The findings and conclusions in this paper are those of the authors, do not necessarily represent the official position of the Centers for Disease Control and Prevention, USAID or the US government, nor do they suggest endorsement of specific products or trade names used in this study.

## Supporting information

## Notes

### Competing Interest Statement

The authors have declared no competing interest.

### Clinical Trial

N|A

### Funding Statement

Yes

### Author Declarations

The survey protocol was approved by the Ethics and Scientific Committee, Ministry of Health and Sanitation, Sierra Leone.

